# A proteogenomic analysis of the adiposity colorectal cancer relationship identifies GREM1 as a probable mediator

**DOI:** 10.1101/2024.02.12.24302712

**Authors:** Matthew A Lee, Charlie A Hatcher, Emma Hazelwood, Lucy J Goudswaard, Konstantinos K Tsilidis, Emma E Vincent, Richard M Martin, Karl Smith-Byrne, Hermann Brenner, Iona Cheng, Sun-Seog Kweon, Loic Le Marchand, Polly A Newcomb, Robert E Schoen, Ulrike Peters, Marc J Gunter, Bethany Van Guelpen, Neil Murphy

## Abstract

Adiposity is an established risk factor for colorectal cancer (CRC). However, the pathways underlying this relationship, and specifically the role of the circulating proteome, is unclear.

Utilizing two-sample Mendelian randomization and colocalization, based on summary data from large sex-combined and sex-specific genetic studies, we estimated the univariable (UV) associations between: (I) adiposity measures (body mass index, BMI; waist hip ratio, WHR) and overall and site-specific (colon, proximal colon, distal colon, and rectal) CRC risk, (II) adiposity measures and plasma proteins, and (III) adiposity-associated plasma proteins and CRC risk. We used multivariable MR (MVMR) to investigate the potential mediating role of adiposity- and CRC-related proteins in the adiposity-CRC association.

BMI and WHR were positively associated with CRC risk, with similar associations by anatomical tumour site. 6,591 adiposity-protein (2,628 unique proteins) and 33 protein-CRC (8 unique proteins) associations were identified using UVMR and colocalization. 1 protein, GREM1 was associated with BMI only and CRC outcomes in a manner that was consistent with a potential mediating role in sex-combined and female-specific analyses. In MVMR, adjusting the BMI-CRC association for GREM1, effect estimates were attenuated - suggestive of a potential mediating role - most strongly for the BMI-overall CRC association in women.

These results highlight the impact of adiposity on the plasma proteome and of adiposity-associated circulating proteins on the risk of CRC. Supported by evidence from *cis*-SNP UVMR and colocalization analyses, GREM1 was identified as a potential mediator of the BMI-CRC association, particularly in women, and warrants further experimental investigation.

## 2. Introduction

Adiposity is an established causal risk factor for the development of colorectal cancer (CRC)^1–3^. However, the underlying biological pathways are not fully understood. Identifying potentially modifiable mediators of the relationship between adiposity and CRC development could uncover targets for pharmacological and/or lifestyle interventions. Circulating proteins are strong candidate mediators. Evidence from molecular epidemiological and genetic studies has linked adiposity with broad changes in the human circulating proteome, including via effects on glucose and lipid metabolism, and inflammatory and immune markers^4–7^. Whether these changes to the proteome influence the association between adiposity and CRC risk is unclear.

Mendelian randomization (MR) uses genetic variants as instrumental variables which, under specific assumptions, can be used to investigate the causal relationship between an exposure and outcome. Given the random allocation of alleles during gametogenesis, across a large enough population, findings of MR analyses are more robust to the effects of confounding and reverse causation than traditional observational studies. Two-step/network MR^8,9^ and Multivariable (MV) MR^10^ analyses can be used to investigate the intermediate effects of traits which may reside on the causal mechanistic pathway from exposure to outcome^11^. To date, MVMR analyses examining the role of the proteome as an intermediate in the relationship between adiposity and CRC have not been undertaken.

Here, we investigated whether circulating proteins act as intermediates in the association between adiposity (BMI and WHR) and CRC risk. We conducted colocalization and MVMR analyses using summary statistics from genome-wide association studies (GWAS) of adiposity traits, circulating protein levels, and CRC risk, and examined if mediating proteins were expressed in adipose and CRC tissue.

## 3. Methods

### 3.1. Study design

Four main analysis steps were performed sequentially (Figure 1) to estimate: (I) the causal relationship between adiposity measures (BMI and WHR) and CRC risk, (II) the causal relationship between adiposity measures and plasma proteins, (III) the causal relationship between proteins and CRC risk, and (IV) the potential mediating effects of adiposity-associated proteins in the adiposity-CRC association. We performed forward and reverse UVMR for steps I-III and used MVMR for step IV. For step III we performed cis-SNP UVMR and colocalization. For all steps, sex-combined and sex-specific analyses were performed. All analyses were performed using R version 4.1.2 and the following packages: TwoSampleMR^12^ (version 0.4.22), MVMR^13^ (version 0.3), and coloc^14^ (version 5.2.0). Forest plots were created using ggforestplot (version 0.1.0).

**Figure 1.**
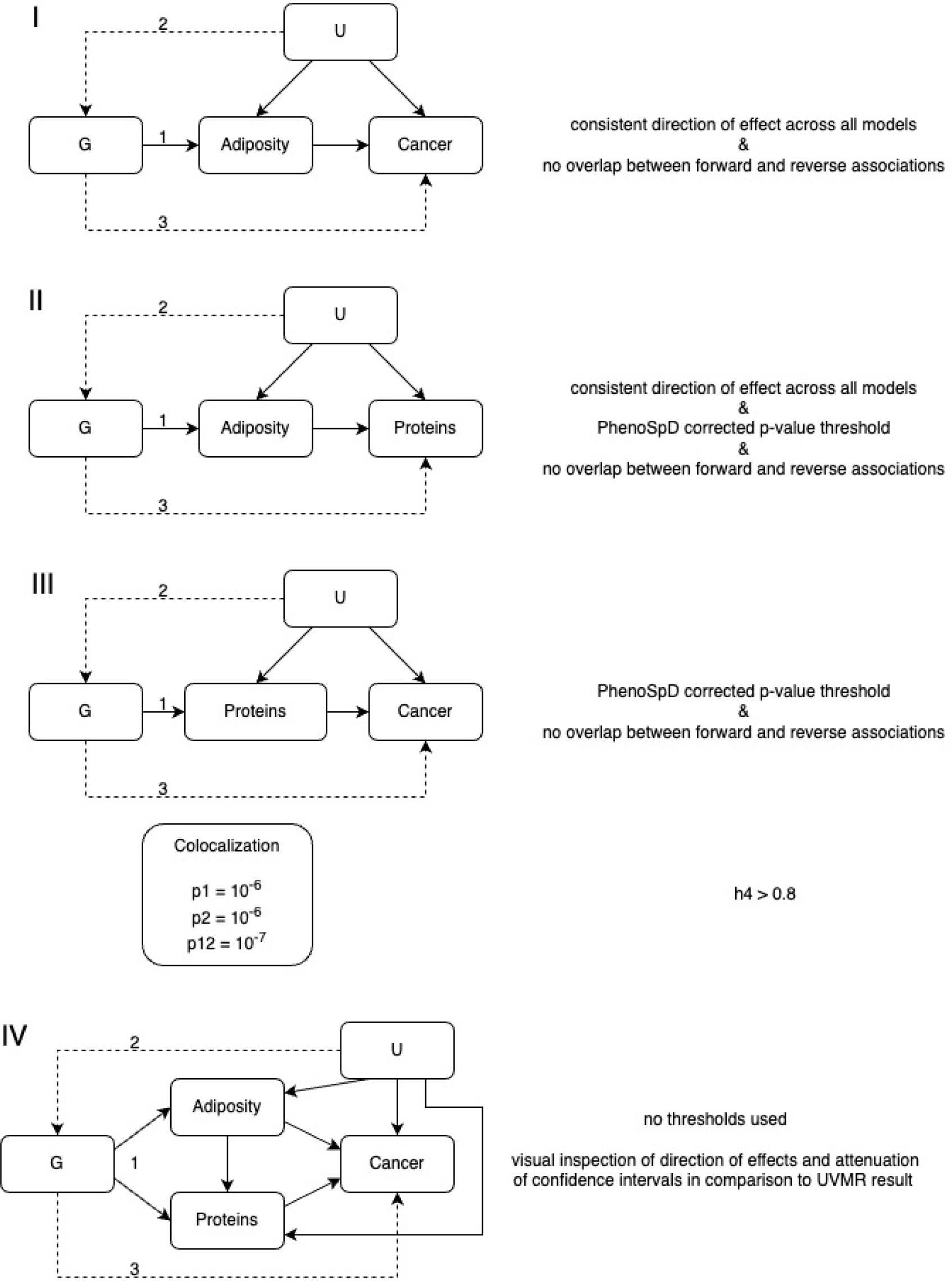
Analysis overview. Directed acyclic graph overview of main analyses: I-III; univariable Mendelian randomization analyses, IV; multivariable MR analysis. Text to the right of each analysis gives the requirements for an association. 1-3: MR assumptions; G: genetic variant(s); U: unmeasured confounders; p*: prior probability of a random SNP in the region (1) being (causally associated with trait 1 and not trait 2, (2) trait 2 and not trait 1, or (12) both traits; h4: probability that there is an association with both traits in the region (shared causal variant).

### 3.2. Data sources and study populations

Details of datasets, study populations, and thresholds used in these analyses are available in Extended Data 1. Briefly, data for adiposity measures were obtained from Pulit et al., (2019)^15^; CRC (overall, colon, proximal, distal, and rectal) from Huyghe et al., (2019)^16^; and up to 4907 plasma proteins from Ferkingstad et al., (2021)^17^. BMI and WHR, available in European ancestries (sex-combined and sex-specific), were calculated as 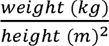 and 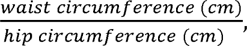 respectively; CRC, available in European and East Asian ancestries (sex-combined and sex-specific), was physician diagnosed; and protein concentrations, available in an Icelandic population (sex-combined), were measured in ethylenediaminetetraacetic acid (EDTA) plasma samples using SomaScan^®^ (SomaLogic, version 4; 4,907 aptamers). The SomaScan platform uses 4,034 modified nucleotides known as Slow Off-rate Modified Aptamers (SOMAmers) which make direct contact with proteins, enabling detection of 3,622 unique proteins or protein complexes and quantifies them in relative fluorescence units (RFUs) using DNA microarray^18^. Separate SOMAmers can bind to isoforms of the same protein but can also bind to the same protein at different sites (which can be impacted by post-translational modifications or complexes formed with other proteins) enabling a larger number of proteins and protein complexes to be quantified.

For all MR analyses, a minimum genome-wide significance threshold of 5 x 10^-8^ was used for all data – more stringent genome-wide significance thresholds were used for adiposity measures and proteins given wider genotyping coverage^19^ – and a linkage disequilibrium (LD) independence threshold of 0.001 was used where applicable (Extended Data 1). For all exposures, F-statistics were calculated for each SNP and a mean was calculated for each instrument, with an F-statistic > 10 indicating a strong instrument^20^. Proteins were included in two UVMR analyses: *cis*- and *trans*-SNPs were used for reverse MR analyses of the association between adiposity measures and proteins to examine reverse causation; cis-SNPs were used in the forward MR analyses of the association between proteins and CRC.

The *cis*-SNPs were obtained directly from the supplementary data of the Ferkingstad et al., paper in which they identified *cis*-SNPs reaching the genome-wide significance threshold (p-value < 1.8 x 10^-9^) as those loci which were ≤1Mb from the transcription start site of the gene encoding the measured protein. All SNPs within a 1Mb region of each *cis*-SNP were merged into a single region, and the SNP with the lowest p-value was considered the ‘sentinel’ variant for that protein. Sentinel variants in LD (r^2^ ≥ 0.8) with one another were considered to be a single ‘pQTL variant’. In total, Ferkingstad et al., identified a single *cis*-SNP for 1,490 of 4,907 proteins.

#### 3.2.1. Units

Data for adiposity and protein measures were inverse rank normally transformed prior to genome-wide analysis. Assuming the distribution of each trait was normal prior to transformation and genome-wide analysis, we interpret these units to be approximately equivalent to a normalized standard deviation (SD) of the respective trait. Estimates and odds ratios (ORs) are therefore interpreted as the change in outcome per normalized SD unit change in the exposure.

### 3.3. Statistical analysis

MR relies upon three core assumptions: (1) the instrument must be associated with the exposure of interest, (2) there are no confounders of the association between the instrument and the outcome, and (3) the instrument is not related to the outcome except via its effect on the exposure of interest. The same assumptions are extended to include the intermediate in MVMR: (1) the instrument must be associated with the exposure given the presence of the mediator, (2) the instrument must be independent of the outcome given both exposure and mediator, and (3) the instrument is not related to the outcome except via its effect on the exposure given the presence of the mediator. Assumption 1 may be satisfied by using a standard genome-wide significance threshold of 5 x 10^-8^ and instruments with an F-statistic, or conditional F-statistic for MVMR analyses^21^, > 10. Assumptions 2 and 3 are unverifiable but were tested using sensitivity models sensitive to the effects of pleiotropy and with *cis*-SNP UVMR and colocalization. Colocalization attempts to differentiate between distinct causal variants and a single shared signal^22^. In combination with UVMR, colocalization can be used to assess the validity of MR assumptions (distinct causal variants likely result from LD) and strengthen evidence for a causal effect.

#### 3.3.1. Identification of associations

An adiposity-CRC association (step I) was identified if there was a consistent direction of effect across all MR models and there was no consistent direction of effect in the reverse MR analyses. The same requirement, plus a PhenoSpD^23–25^ corrected p-value threshold (we used the more conservative of the two approaches applied by PhenoSpD), were used to identify adiposity-protein associations (step II). Only proteins with cis-SNP information were used in the subsequent UVMR and MVMR analyses. A protein-CRC association (step III) was identified if the PhenoSpD corrected p-value threshold was met, there was no consistent direction of effect across all MR models in the reverse MR, and evidence of colocalization (h4 ≥ 0.8) was observed. We interpreted a protein as having a potential mediating role in the adiposity and CRC relationship if the MVMR result adjusting for that protein attenuated towards the null when compared with the UVMR result (step IV).

#### 3.3.2. Univariable Mendelian randomization

For all exposures, the following summary-level data were obtained from the original GWAS: rsID, effect allele, other/non-effect allele, effect allele frequency, effect estimate, standard error of the effect estimate, p-value, sample size, and units. Genetic variants were extracted from each outcome GWAS and, where these were not present, proxy SNPs were included if LD was ≥ 0.8. For proxy SNPs, the inclusion of SNPs where the reference strand was ambiguous (strand flips) was allowed and the reference strand was inferred using a minor allele frequency (MAF) threshold so long as the MAF was not ≥ 0.3, in which case the proxy SNP was excluded. Exposure and outcome summary statistics for each of the exposure-related SNPs were harmonised in reference to the exposure effect allele being on the increasing scale. For included alleles where the reference strand was ambiguous, the positive strand was inferred using effect allele frequency (EAF) so long as the EAF was not within 0.3 – 0.7, otherwise the SNP was removed.

An inverse variance weighted (IVW), multiplicative random effects (IVW-MRE) model was used to estimate the effect of each exposure on the outcome. The model assumes that the strength of the association of the genetic instruments with the exposure is not correlated with the magnitude of the pleiotropic effects and that the pleiotropic effects have an average value of zero^26^. Where only one SNP was present, the Wald ratio was used (e.g., *cis*-SNP UVMR).

##### 3.3.2.1. Sensitivity analysis

The assumptions of no pleiotropy among genetic instruments and outcomes were explored using: MR-Egger^27^, weighted median^28^, and weighted mode^29^ based estimators where ≥ 3 SNPs were available. MR-Egger provides an estimate of unbalanced or directional horizontal pleiotropy via the intercept of a linear regression of the SNP-exposure and SNP-outcome association^27^. The weighted median provides consistent estimates when at least 50% of included instruments are invalid^28^. The weighted mode assumes the true causal effect is the most common effect and it is robust when most effect estimates are derived from valid instruments^29^. In addition, a single-SNP MR using the Wald ratio and a “leave-one-out” MR sensitivity analysis were conducted to assess the influence of individual variants on the observed associations.

#### 3.3.3. Colocalization

For each protein, the sentinel cis-SNP identified by Ferkingstad et al., was extracted along with a 1Mb window. This region was then extracted from each CRC GWAS and colocalization was implemented using the single causal variant approach of Giambartolomei et al., (2014)^30^. The LD matrix was generated using the 1000 genomes reference panel (phase 3) and priors were set at p^1^ = 10^-6^, p^2^ = 10^-6^, and p^12^ = 10^-7^ based on a window of 5,000 SNPs using https://chr1swallace.shinyapps.io/coloc-priors/ (accessed 15/05/2023).

#### 3.3.4. Multivariable Mendelian randomization

SNPs associated with the exposure (adiposity) and proposed intermediate proteins (*cis*- and *trans*-SNPs) were extracted and combined. This combined SNP list was extracted from the adiposity GWAS. The resulting instrument was then clumped to remove duplicate SNPs and SNPs in LD with one another using the same clumping thresholds as with the UVMR analysis. An IVW model was used to obtain the direct causal effect of each adiposity measure adjusted for each protein on CRC risk. Instrument strength for each exposure was estimated using a generalized version of Cochran’s Q^31^ assuming a pairwise covariance of zero^13^.

#### 3.3.5. Protein expression analyses

To investigate whether proteins included in the MVMR analyses were expressed in adipose and CRC tissue we used GTEx^32^ (version 8) data to compare protein tissue expression relative to whole blood using the Wilcoxon rank sum test and visualised expression profiles using violin plots.

## 4. Results

An overview of the datasets used, protein information, and all results from the MR and colocalization analyses are presented in Extended Data 1. PhenoSpD identified a total of 1,293 independent variables, and a PhenoSpD corrected p-value threshold of 3.97 x 10^-5^ was, therefore used (Extended Data 2).

### 4.1. Association between adiposity measures and colorectal cancer

In step I, BMI and WHR were positively associated with overall and site-specific CRC risk in men and women (Figure 2). Sensitivity models were broadly consistent (directions of effect estimates were in the same direction as the IVW-MRE model and CIs overlapped) with the main IVW-MRE model. However, inconsistent directions were observed for the association between WHR and distal colon cancer and overall CRC in men and rectal cancer in women (Extended data 3). Furthermore, the reverse UVMR analyses (Extended data 4) showed an increasing effect of proximal and distal colon cancer on WHR. In both cases, sensitivity analyses produced generally consistent results, particularly for proximal colon cancer in men and for distal colon cancer in women. These pairings were, therefore, excluded from subsequent MVMR analyses.

**Figure 2.**
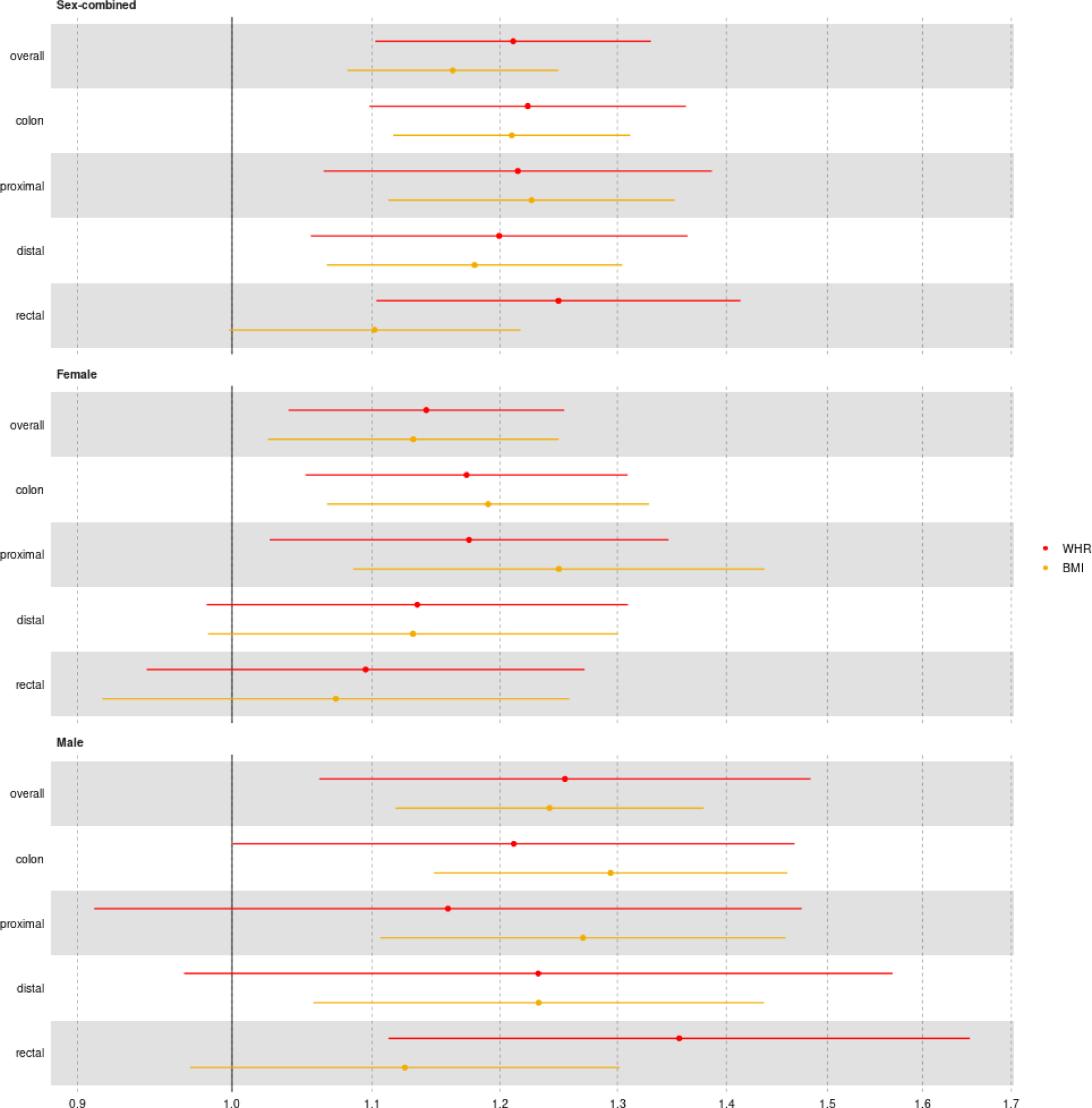
Association between adiposity measures and colorectal cancer outcomes. Odds ratios and 95% confidence intervals shown for the main analysis using the inverse variance weighted multiplicative random effects (IVW-MRE). BMI = body mass index; WHR = waist hip ratio

### 4.2. Association between adiposity measures and proteins

In step II, 6,591 adiposity-protein (2,628 unique proteins) associations were identified (consistent directions of effect across MR models, PhenoSpD corrected p-value threshold reached, and no conflicting association identified in the reverse UVMR; Figure 3) across the sex-combined and sex-specific analyses. The largest number of associations were identified for the female-specific analysis of BMI (N = 2,395 BMI-associated proteins), with more analyses reaching the PhenoSpD-corrected p-value threshold in the female analysis compared to the male analysis (Figure 4).

**Figure 3.**
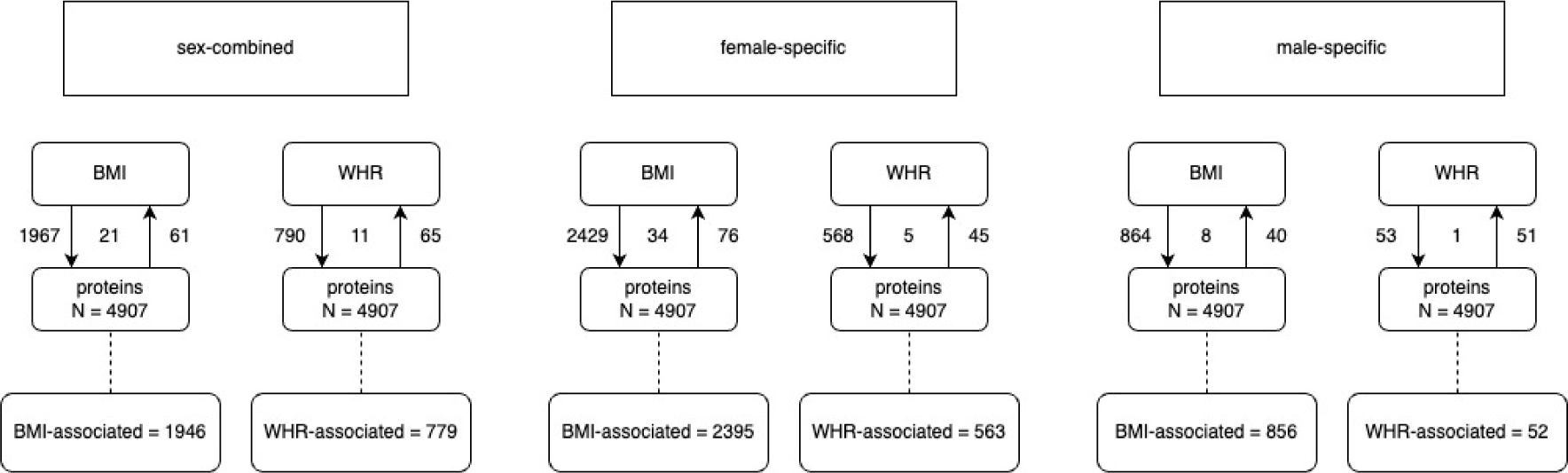
Overview of associations between adiposity measures and plasma proteins (step II in the main analysis plan). Arrows show the direction of the univariable Mendelian randomization (UVMR) analysis. Values on the outside of the lines indicate the number of associations identified in that direction; values in between the lines indicate the number of associations identified in both directions and for which there is, therefore, conflicting evidence of association. N = gives the number of proteins available for analysis. BMI-WHR-associated gives the number of proteins associated with the adiposity measure. BMI = body mass index; WHR = waist hip ratio.

**Figure 4.**
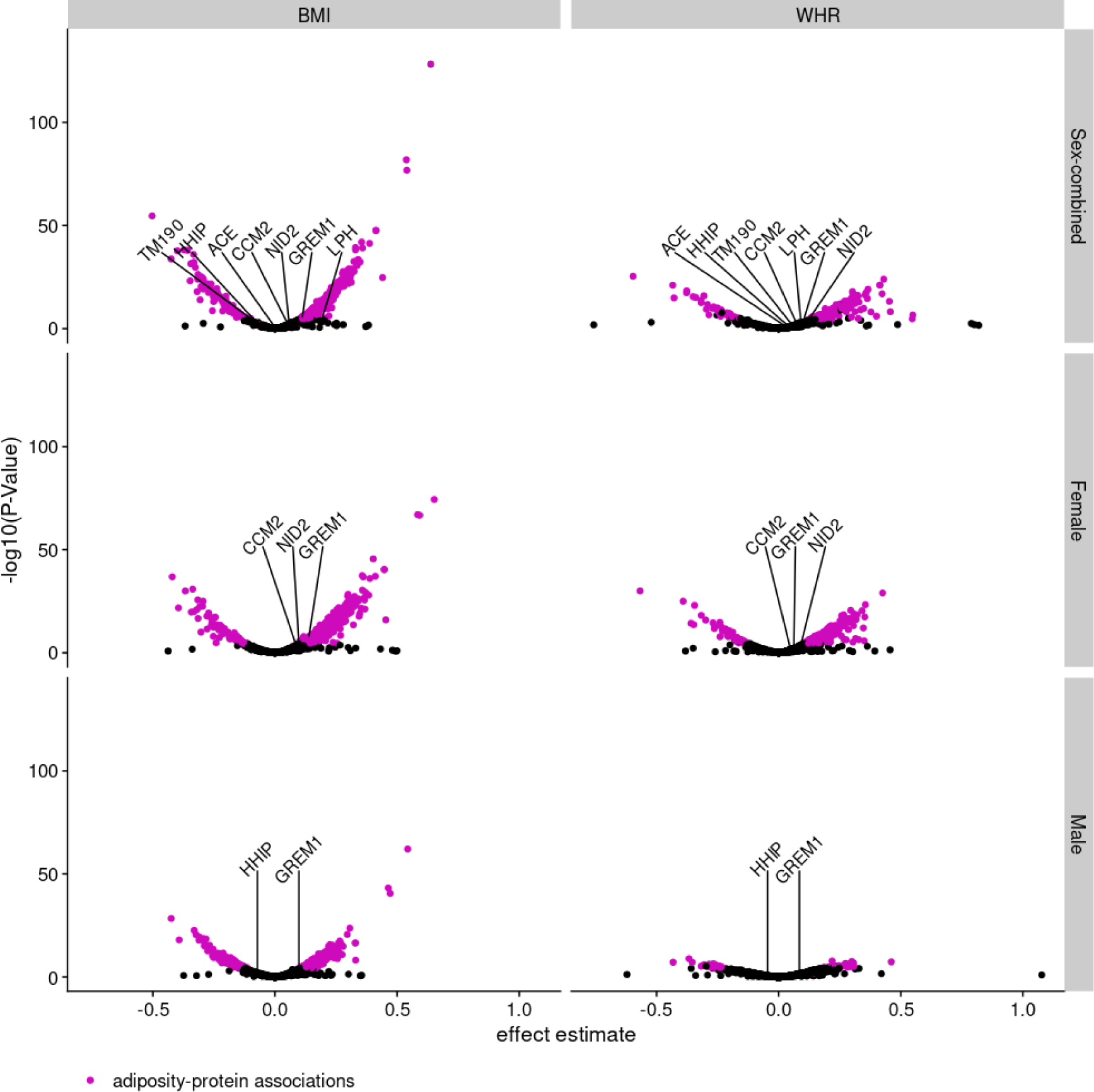
Association between adiposity measures and proteins in step II of the main analysis plan. The volcano plot shows effect estimates and -log10(pval). Adiposity-protein associations are highlighted in purple (analyses reaching the PhenoSpD corrected p-value (0.05/1293), consistent directions of effect across MR models, and no conflicting association identified in the reverse UVMR). Protein labels highlight those proteins which were associated with colorectal cancer outcomes in the UVMR analysis in step III of the main analysis plan. BMI = body mass index; WHR = waist hip ratio.

### 4.3. Association between proteins and colorectal cancer

In step III, it was possible to perform UVMR analysis of up to 962 proteins in relation to CRC outcomes using *cis*-SNPs. In total, 35 protein-CRC (8 unique proteins) associations were identified (PhenoSpD-corrected p-value threshold reached and no conflicting association identified in the reverse UVMR; Figure 5) across the sex-combined and sex-specific UVMR analyses. In the male-specific analyses, 65 reverse associations were identified between proteins and distal colon cancer risk. However, there were no conflicts with the forward UVMR analysis in this or any other protein-CRC analyses. There was evidence (h4 ≥ 0.8) of colocalization for 87 protein-CRC pairs (25 unique proteins) across the sex-combined and sex-specific analyses of the CRC outcomes (Extended Data 4). Of the 35 protein-CRC UVMR associations, 2 were not corroborated by colocalization (h4 < 0.5 for the protein GRFAL in relation to overall CRC and rectal cancer risk in sex-combined analysis).

**Figure 5.**
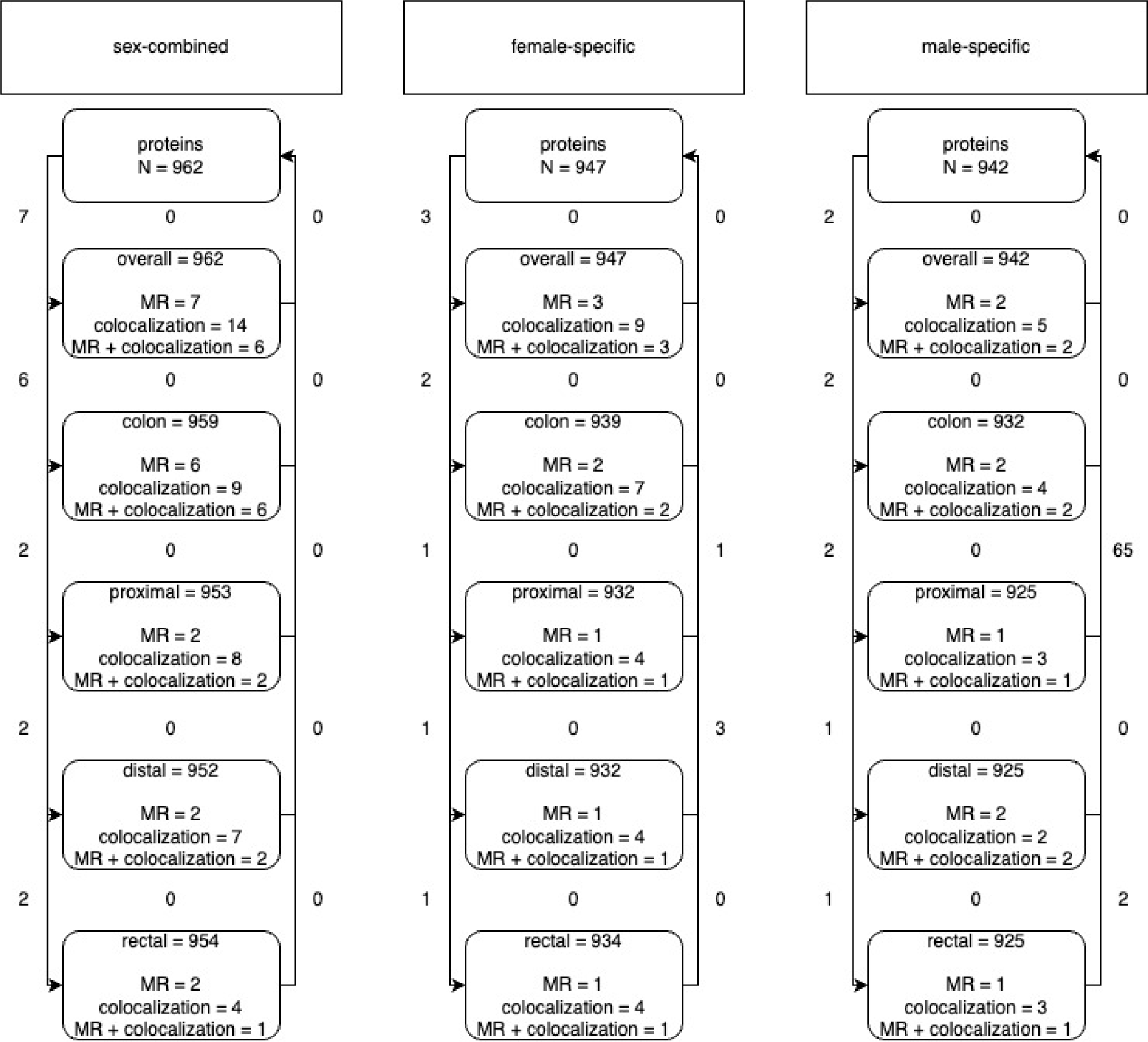
Overview of associations between proteins and colorectal cancer (CRC) outcomes (step III in the main analysis plan). Arrows show the direction of the univariable Mendelian randomization (UVMR) analysis. Values on the outside of the lines indicate the number of associations identified in that direction; values in between the lines indicate the number of associations identified in both directions and for which there is, therefore, conflicting evidence of association. N = gives the number of proteins available for analysis. MR = gives the number of cis-SNP UVMR analyses which reached the PhenoSpD pvalue threshold for that analysis. Colocalization = gives the number of proteins which colocalized with that CRC outcome. MR + colocalization = gives the overlap between the cis-SNP UVMR and colocalization analyses and indicates the protein-CRC associations.

**Figure 6.**
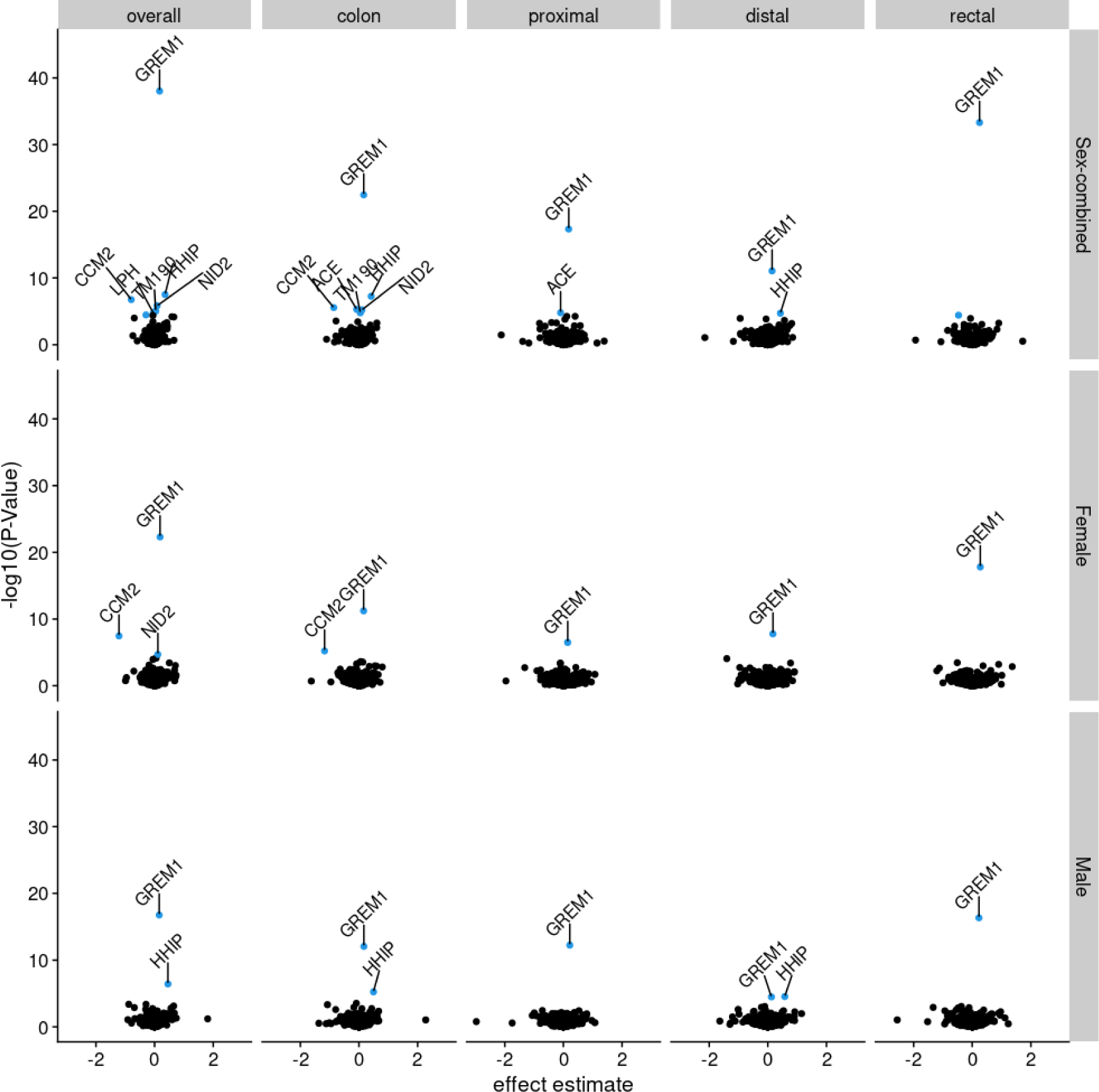
Association between adiposity-related proteins and colorectal cancer (CRC) outcomes in step III of the main analysis plan. The volcano plot shows effect estimates and -log10(pval) with analyses reaching the PhenoSpD corrected p-value (0.05/1293) highlighted in blue and analyses reaching the PhenoSpD corrected-pvalue and with evidence of colocalization labelled with the protein name. The X-axis has been constrained to −3 – 3, excluding 3 analyses which did not meet any association thresholds: PTP4A2 and proximal colon cancer in males (effect estimate = 102) and NANS and distal colon cancer in males (effect estimate = 19) and females (effect estimate = 19).

### 4.4. Multivariable Mendelian randomization

Of the 7 proteins associated with CRC outcomes across the sex-combined and sex-specific analyses, the results for only 1 (GREM1) were directionally consistent with a possible mediating role of the association between adiposity and CRC risk. That is, an increase in BMI was associated with an increase in GREM1 and an increase in GREM1 was associated with an increase in CRC risk. In MVMR (step IV) we considered a protein as having a potential mediating role if, in comparison to the UVMR estimate of the association between adiposity and CRC, the adjusted estimate was attenuated (i.e., the effect estimate tends towards the null and the CI overlaps the null). For GREM1, effect estimates and CIs for all MVMR analyses tended towards the null. However, evidence of attenuation was observed solely for the female-specific analysis of overall CRC risk (Figure 7). Attenuation of the sex-combined analysis of rectal CRC was also observed, but the CI of the UVMR effect of BMI on rectal cancer risk also overlapped the null. Conditional F-statistics were > 10 (Extended Data 4). There was evidence that all but the female-specific analysis of distal colon cancer used invalid instruments (Q statistic p-value < 0.05).

**Figure 7.**
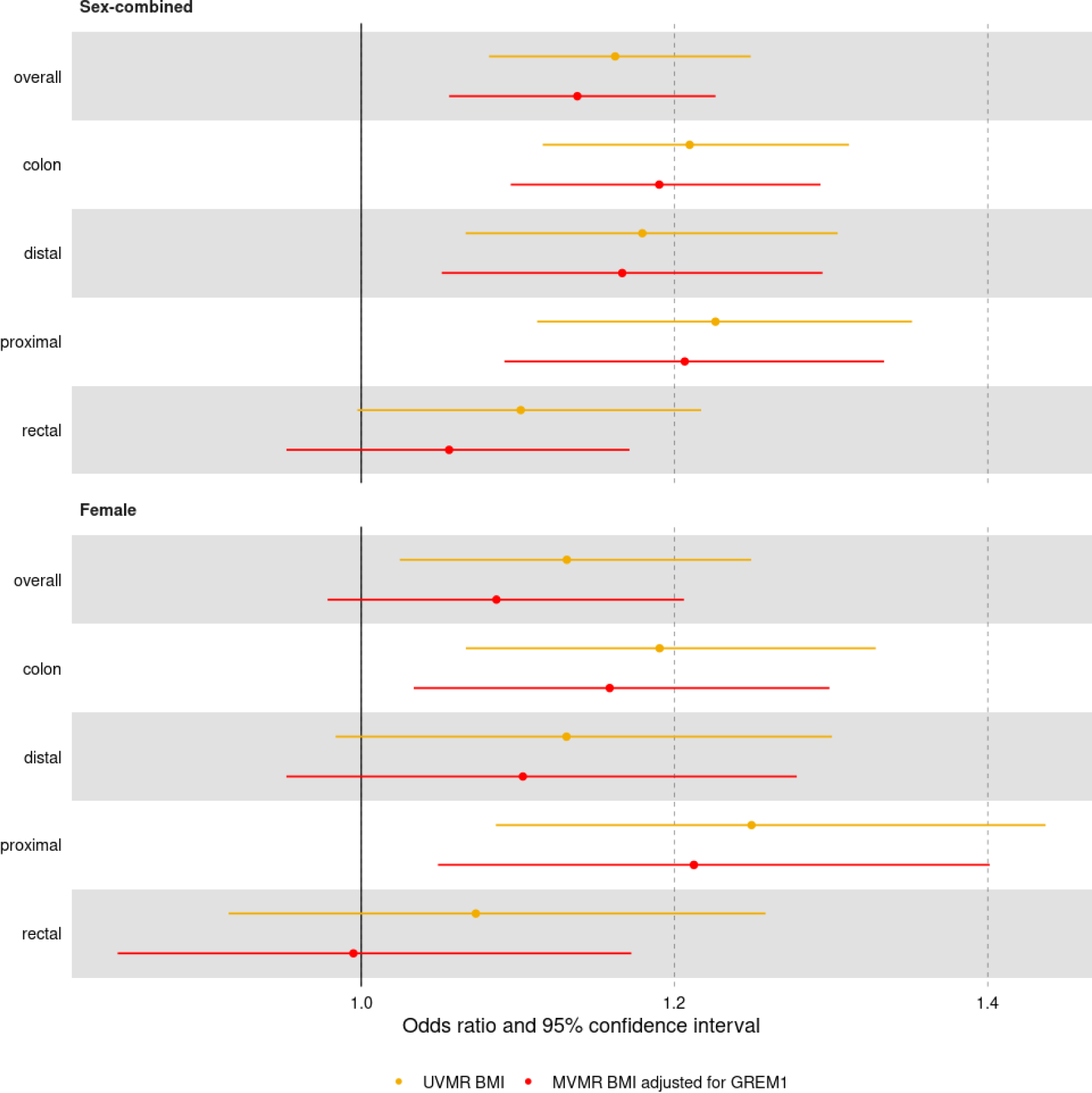
Association between body mass index (BMI) and colorectal cancer outcomes using univariable (UV; orange line) and multivariable (MV; red lines) Mendelian randomization (MR). In these MVMR analyses (step IV), the effect of BMI on colorectal cancer outcomes is estimated after adjusting for the effect of GREM1. Odds ratios for the inverse variance weighted multiplicative random effects model shown alongside 95% confidence intervals. No adiposity-protein-CRC associations were identified in the male UVMR analyses and as such MVMR was not performed.

### 4.5. Follow-up analysis

Using GTEx data, GREM1 was found to be differentially expressed (Bonferroni corrected p-value = 0.05/53) in most tissues compared to whole blood, with some of the highest levels noted in the gastrointestinal tract and visceral adipose tissue in both sexes (Figure 8).

**Figure 8.**
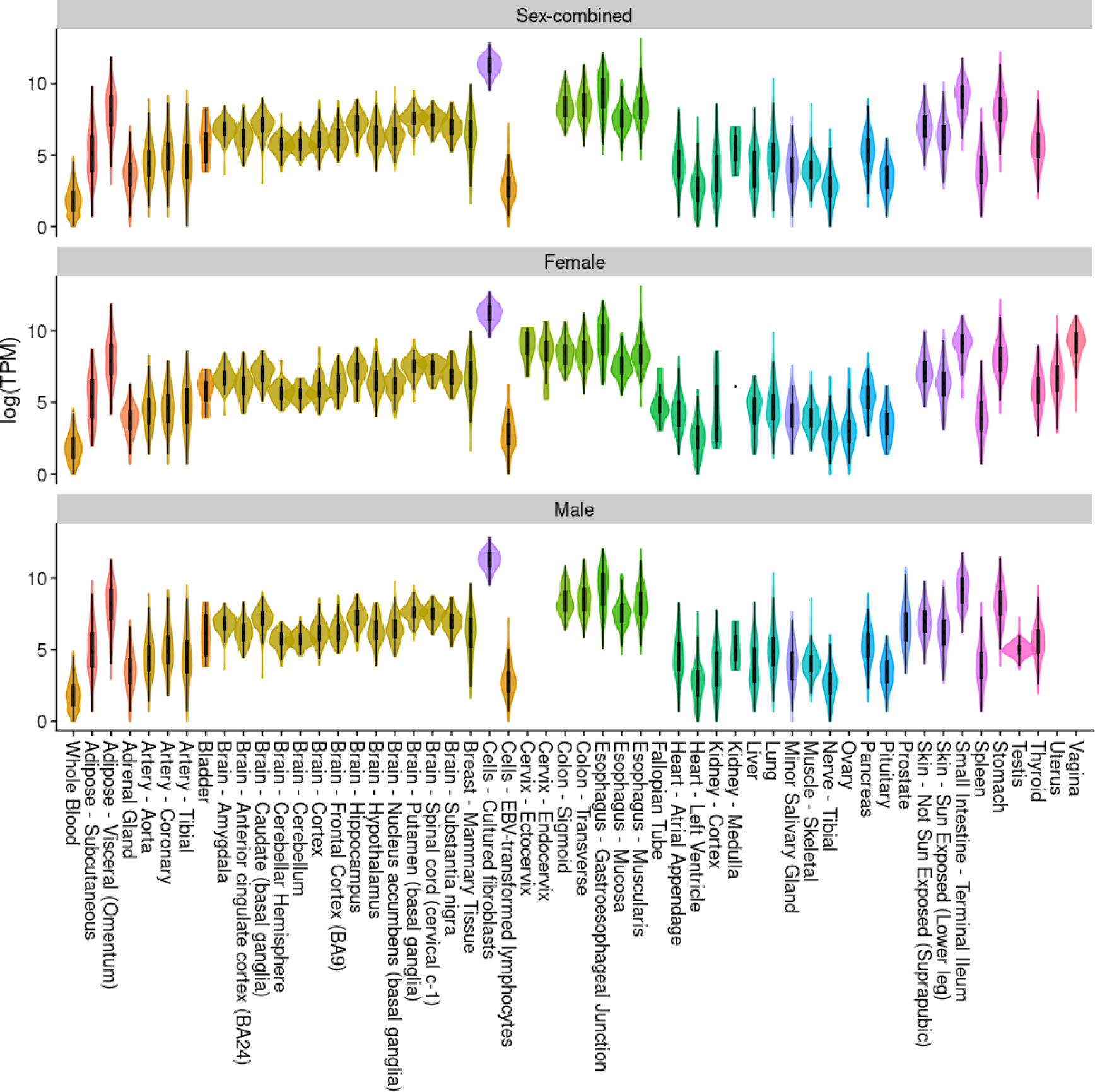
Tissue gene expression profile of GREM1. The violin plot presents expression levels as log transcripts per million (TPM). Data are from GTEx version 8^32^. Box plots are shown with the interquartile range (25^th^ and 75^th^ percentiles). Colours distinguish different tissue types (e.g., adipose and brain).

## 5. Discussion

Using complementary MR and colocalization analyses we conducted the largest and most comprehensive study to date examining the role of the plasma proteome as an intermediate in the relationship between adiposity and CRC development. We found novel evidence of a potential mediating role of the protein GREM1, primarily in the association between BMI and overall CRC risk in women. For the remainder of our analyses, we found little evidence of individual proteins acting as intermediates in the adiposity-CRC relationship.

GREM1, a bone morphogenic protein (BMP) antagonist^33^, has previously been linked with CRC^16,17,34,35^, and is expressed in many tissues, including adipose and colon tissue. BMPs play an important role in embryonic development and morphogenesis and may be considered tumour repressors as inhibition of BMP signalling has been associated with a number of cancers^36,37^. GREM1 is associated with proliferation, angiogenesis, and epithelial-to-mesenchymal transition of cancer cells^36^ and may be involved in colon cancer tumour progression^38^, with a number of studies linking GREM1 to CRC development^34,35,39^. In addition, a recent gene-environment interaction analysis identified a locus in the *FMN1*/*GREM1* gene region that interacted with BMI on the positive association with CRC risk^40^. The exact mechanisms underlying the GREM1-CRC relationship are unclear but may be related to expression in the tumour microenvironment given that GREM1 expression is lower in CRC tissue than adjacent non-cancerous and normal tissue^34^. Increased GREM1 expression in CRC tissue has also been associated with low tumour stage and a more favourable prognosis^41^, and increased GREM1 expression is found in the tumour microenvironment, such as in visceral adipose tissue^42^ and colonic crypt bases via cancer associated fibroblasts^43^.

To our knowledge, this was the first comprehensive proteogenomic analysis conducted so far to estimate the role of the plasma proteome as an intermediate of the positive association found between adiposity and CRC. Our study has several additional notable strengths. Adiposity-protein-CRC triples included in the MVMR analyses were supported by consistent directions of effect across the main IVW-MRE model and sensitivity models (MR-Egger, weighted median, and weighted mode). We also conducted reverse UVMR analyses for all adiposity-CRC, adiposity-protein, and protein-CRC analyses to identify possible reverse causation and exclude pairs where evidence was conflicting. Several limitations should also be acknowledged. Although the availability of sex-specific summary statistics from large genome-wide association studies were available for adiposity measures and CRC, only sex-combined protein data were available which may have led to biased estimates, especially for some of the MVMR analyses, in which the adiposity exposure and outcome were both sex-specific. Instrument strength, measured via F-statistics, was appropriate for most analyses, including the MVMR analyses. However, there was evidence of weak instrument bias across most MVMR analyses which can lead to estimates for the exposure and intermediate moving towards and away from the null^13^. In addition, *cis*-SNPs identified by Ferkingstad *et al*., were available for less than half of the 4,907 proteins included in the adiposity-protein UVMR analysis.

Our results highlight the broad impact of adiposity on the plasma proteome and of adiposity-associated circulating proteins on the risk of developing CRC. We found robust evidence of a causal effect of one adiposity-related protein, GREM1, as a likely mediator of the adiposity and CRC relationship, particularly for women. Given evidence from previous studies highlighting the relationships between GREM1 and CRC, our results suggest that the GREM1 pathway may be a potential mechanism underlying the obesity-CRC relationship and future experimental investigation is warranted.

## 6. Author contributions

MAL: Data curation, Formal analysis, Investigation, Methodology, Project administration, Visualisation, Writing – original draft, Writing – review and editing

CAH: Methodology, Writing – review and editing

EH: Methodology, Writing – review and editing

LG: Writing – review and editing

UP: Writing – review and editing

KKT: Writing – review and editing

EEV: Writing – review and editing

RMM: Writing – review and editing

MJG: Conceptualization, Funding acquisition, Methodology, Project administration, Resources, Supervision, Writing – review and editing

HB: Writing – review and editing

IC: Writing – review and editing

SK: Writing – review and editing

LLM: Writing – review and editing

PAN: Writing – review and editing

RES: Writing – review and editing

BVG: Conceptualization, Funding acquisition, Methodology, Project administration, Resources, Supervision, Writing – review and editing

NM: Conceptualization, Funding acquisition, Methodology, Project administration, Resources, Supervision, Writing – review and editing

## 8. Acknowledgements & Funding

Funding (IIG_FULL_2021_030) was obtained from Wereld Kanker Onderzoek Fonds (WKOF), as part of the World Cancer Research Fund International grant programme, and Cancer Research UK (C18281/A29019). NM is supported by the French National Cancer Institute (INCa SHSESP20, grant No. 2020-076). BVG is supported by the Swedish Cancer Society (fellowship No. 21 0467 FE 01 H and project grant No. 20 1154 PjF). EH is supported by a Cancer Research UK Population Research Committee Studentship (C18281/A30905 and is part of the Medical Research Council Integrative Epidemiology Unit at the University of Bristol which is supported by the Medical Research Council (MC_UU_00032/03) and the University of Bristol. LJG is supported by a Transition Fellowship as part of the British Heart Foundation Accelerator Award (AA/18/1/34219) and an Academic Career Development Fund (University of Bristol). RMM is a National Institute for Health Research Senior Investigator (NIHR202411). RMM is supported by a Cancer Research UK 25 (C18281/A29019) programme grant (the Integrative Cancer Epidemiology Programme). RMM is also supported by the NIHR Bristol Biomedical Research Centre which is funded by the NIHR (BRC-1215-20011) and is a partnership between University Hospitals Bristol and Weston NHS Foundation Trust and the University of Bristol. RMM is affiliated with the Medical Research Council Integrative Epidemiology Unit at the University of Bristol which is supported by the Medical Research Council (MC_UU_00011/1, MC_UU_00011/3, MC_UU_00011/6, and MC_UU_00011/4) and the University of Bristol. Department of Health and Social Care disclaimer: The views expressed are those of the author(s) and not necessarily those of the NHS, the NIHR or the Department of Health and Social Care.

## 9. Disclaimer

Where authors are identified as personnel of the International Agency for Research on Cancer/World Health Organization, the authors alone are responsible for the views expressed in this article and they do not necessarily represent the decisions, policy or views of the International Agency for Research on Cancer/World Health Organization.

## 10. Data availability

### 10.1. Underlying data

This work is supported by a GitHub repository (https://github.com/mattlee821/adiposity_proteins_colorectal_cancer) which is archived on Zenodo (https://zenodo.org/record/7780822#.ZCQ3U-xBz0o). Here, all publicly available data, code, and results used in this work are available. The full summary statistics for BMI and WHR are publicly available from Zenodo (https://zenodo.org/record/1251813#.Yk7O25PMIUE). The full summary statistics for all proteins are publicly available from DECODE (https://download.decode.is/form/folder/proteomics). The full summary statistics for CRC are not publicly available but can be obtained from GECCO (https://www.fredhutch.org/en/research/divisions/public-health-sciences-division/research/cancer-prevention/genetics-epidemiology-colorectal-cancer-consortium-gecco.html).

### 10.2. Extended Data

This project contains the following extended data available from Zenodo https://zenodo.org/record/7780822#.ZCQ3U-xBz0o):

#### Extended data 1 is an Excel file of 8 tables

1. Overview of all data used, including doi of source. Columns: N = overall sample size or case sample size; pvalue_threshld = genome-wide significance threshold used to identify associated single nucleotide polymorphisms (SNPs); LD_r2 = linkage disequilibrium independence threshold; LD_window = LD independence base window; SNPs = associated SNPs identified using pvalue and LD thresholds; f_stat = mean f-statistics for the associated SNPs; measure = how the trait was measured in the source; adjustment = how the trait was adjusted during genome-wide analysis; transformation = transformation of the trait prior to genome-wide analysis; unit = how the trait is expressed given the transformation.

2. Extended information on proteins sourced from Ferkingstad et al.

3. All publicly available exposure data used in Mendelian randomization analyses – this does not include colorectal cancer data. Columns: CHR = chromosome; POS = base position; SNP = rsID; eaf.exposure = effect allele frequency; beta.exposure = effect estimate; se.exposure = standard error of the beta; pval.exposure = pvalue of the effect estimate and standard error.

4-8. All results produced in the Mendelian randomization (MR) and colocalization analyses, split by analysis: adiposity-cancer, adiposity-protein, protein-cancer, colocalization, multivariable MR. Columns: nsnp = number of SNPs used for the exposure instrument; b = effect estimate; se = standard error; pval = pvalue of the effect estimate and standard error; forward always refers to column 1 as the exposure and column 2 as the outcome; reverse always refers to column 2 as the exposure and column 1 as the outcome; Qstat/Qpval = Cochranes Q statistic and associated pvalue; fstat_adiposity/fstat_protein = the F-statistic associated with adiposity and protein instrument used in the analysis; nsnp_colocalization = number of SNPs included in the 1Mb window used for colocalization; h0-4 = hypothesis priors; h0 = neither trait has a genetic association in the region; h1 = only trait 1 has a genetic association in the region; h2 = only trait 2 has a genetic association in the region; h3 = both traits are associated, but with different causal variants; h4 = both traits are associated and share a single causal variant.

Extended data 2. PhenoSpD results. File gives the eigenvalue associated with each factor in the correlation matrix and the variance of these values. We use the effective number of independent variables calculated using the method of Li and Ji (2005)^25^.

Extended data 3. Association between adiposity measures and colorectal cancer outcomes. Effect estimates and 95% confidence intervals shown for the main analysis using the inverse variance weighted multiplicative random effects (IVW-MRE) model and 3 sensitivity models. BMI = body mass index; WHR = waist hip ratio.

Extended data 4. Association between colorectal cancer measures and adiposity measures. Effect estimates and 95% confidence intervals shown for the main analysis using the inverse variance weighted multiplicative random effects (IVW-MRE) model and 3 sensitivity models. BMI = body mass index; WHR = waist hip ratio.

*Extended data 5. STROBE-MR checklist*^44,45^.

