## Supplementary figures and images for "A proteogenomic analysis of the adiposity colorectal cancer relationship identifies GREM1 as a probable mediator"

### Extended Data 3

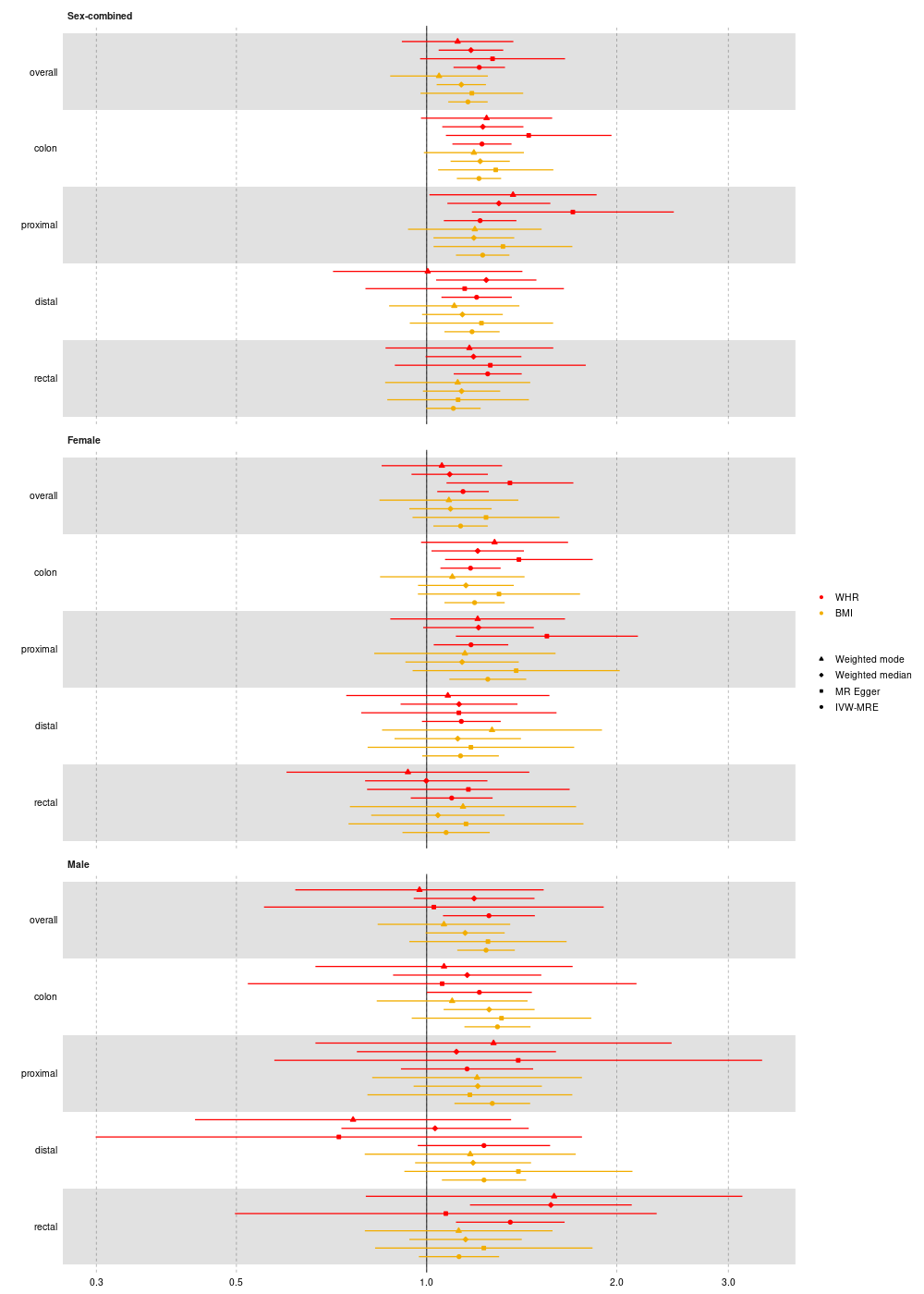

### Extended Data 4

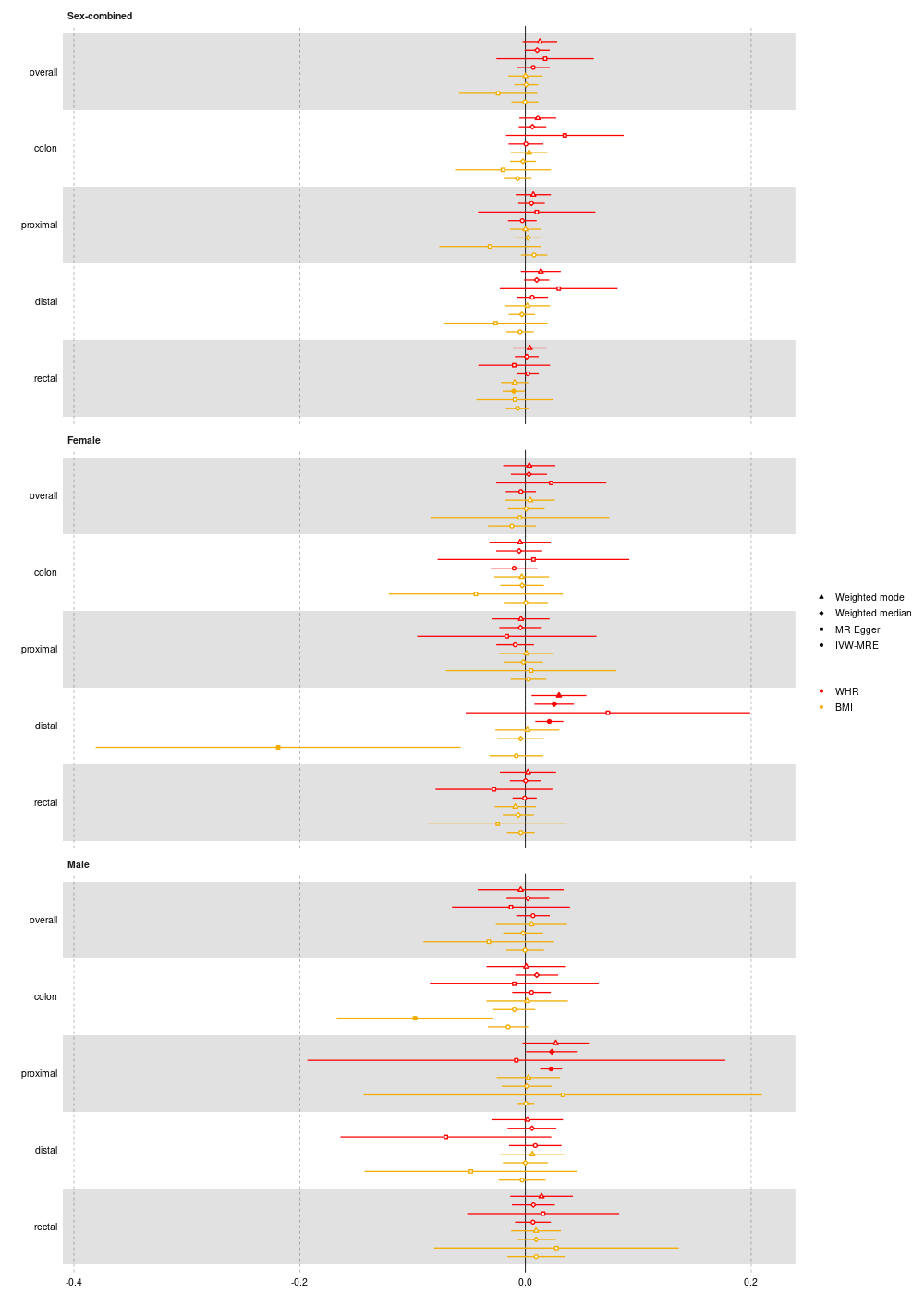
