## Extended Data 5 for "A proteogenomic analysis of the adiposity colorectal cancer relationship identifies GREM1 as a probable mediator"

**STROBE-MR checklist of recommended items to address in reports of Mendelian randomization studies**^1^ ^2^

| **Item No.** | **Section** | **Checklist item** | **Page No.** | **Relevant text from manuscript** |
| --- | --- | --- | --- | --- |
| 1 | **TITLE and ABSTRACT** | Indicate Mendelian randomization (MR) as the study’s design in the title and/or the abstract if that is a main purpose of the study | 1-2 |  |
|  | **INTRODUCTION** |  |  |  |
| 2 | **Background** | Explain the scientific background and rationale for the reported study. What is the exposure? Is a potential causal relationship between exposure and outcome plausible? Justify why MR is a helpful method to address the study question | 3 | …the biological pathways that underlie these relationships are not fully understood. Emerging evidence from observational and genetic studies has linked adiposity with broad changes in the human circulating proteome…  Two-step/network MR^8,9^ and Multivariable (MV) MR^10^ analyses can be used to investigate the intermediate effects of traits which may reside on the pathway from exposure to outcome^11^…  Here, we investigated whether circulating proteins act as intermediates in the positive association between adiposity (BMI and WHR) and CRC. |
| 3 | **Objectives** | State specific objectives clearly, including pre-specified causal hypotheses (if any). State that MR is a method that, under specific assumptions, intends to estimate causal effects |  | …Whether these changes to the proteome influence the obesity and CRC association is unclear.  Mendelian randomization (MR) uses genetic variants as instrumental variables which, under specific assumptions, can be used to investigate the causal relationship between an exposure and outcome. |
|  | **METHODS** |  |  |  |
| 4 | **Study design and data sources** | Present key elements of the study design early in the article. Consider including a table listing sources of data for all phases of the study. For each data source contributing to the analysis, describe the following: |  |  |
|  | a) | Setting: Describe the study design and the underlying population, if possible. Describe the setting, locations, and relevant dates, including periods of recruitment, exposure, follow-up, and data collection, when available. | 4-7 & Extended Data 1 and 2 | Four main analyses were performed sequentially (Figure 1) to estimate: (I) the association between adiposity measures (BMI and WHR) and CRC, (II) the association between adiposity measures and plasma proteins, (III) the association between adiposity-associated plasma proteins (identified in step II) and CRC, and (IV) the potential mediating effect of adiposity-associated plasma proteins in the adiposity CRC association (identified in step II and III). |
|  | b) | Participants: Give the eligibility criteria, and the sources and methods of selection of participants. Report the sample size, and whether any power or sample size calculations were carried out prior to the main analysis | 4-7 & Extended Data 1 and 2 |  |
|  | c) | Describe measurement, quality control and selection of genetic variants | 8,9 |  |
|  | d) | For each exposure, outcome, and other relevant variables, describe methods of assessment and diagnostic criteria for diseases | 6,7 Extended Data 1 and 2 |  |
|  | e) | Provide details of ethics committee approval and participant informed consent, if relevant | NA |  |
| 5 | **Assumptions** | Explicitly state the three core IV assumptions for the main analysis (relevance, independence and exclusion restriction) as well assumptions for any additional or sensitivity analysis | 8 | MR relies upon 3 core assumptions (illustrated in Figure 1)… |
| 6 | **Statistical methods: main analysis** | Describe statistical methods and statistics used |  |  |
|  | a) | Describe how quantitative variables were handled in the analyses (i.e., scale, units, model) | 7 |  |
|  | b) | Describe how genetic variants were handled in the analyses and, if applicable, how their weights were selected | 8 |  |
|  | c) | Describe the MR estimator (e.g. two-stage least squares, Wald ratio) and related statistics. Detail the included covariates and, in case of two-sample MR, whether the same covariate set was used for adjustment in the two samples | 8 Extended Data 1 and 2 |  |
|  | d) | Explain how missing data were addressed | NA |  |
|  | e) | If applicable, indicate how multiple testing was addressed | 8,9 Figure 1 |  |
| 7 | **Assessment of assumptions** | Describe any methods or prior knowledge used to assess the assumptions or justify their validity | 8,10,11 | F-statistic, conditional F-statistic, genome-wide significance thresholds appropriate for genotyping coverage, MR sensitivity models (e.g., Egger), cis-SNP UVMR, colocalization |
| 8 | **Sensitivity analyses and additional analyses** | Describe any sensitivity analyses or additional analyses performed (e.g. comparison of effect estimates from different approaches, independent replication, bias analytic techniques, validation of instruments, simulations) | 11 | cis-SNP UVMR, colocalization |
| 9 | **Software and pre-registration** |  |  |  |
|  | a) | Name statistical software and package(s), including version and settings used | 4 | All analyses were performed using R version 4.1.2 and the following packages TwoSampleMR^13^ (version 0.4.22), MVMR^14^ (version 0.3), and coloc^15^ (version 5.2.0), forestplots were created using ggforestplot (version 0.1.0). |
|  | b) | State whether the study protocol and details were pre-registered (as well as when and where) | NA | Study was not pre-registered |
|  | **RESULTS** |  |  |  |
| 10 | **Descriptive data** |  |  |  |
|  | a) | Report the numbers of individuals at each stage of included studies and reasons for exclusion. Consider use of a flow diagram | Extended data 1 and 2 |  |
|  | b) | Report summary statistics for phenotypic exposure(s), outcome(s), and other relevant variables (e.g. means, SDs, proportions) | Extended data 1,2,3 |  |
|  | c) | If the data sources include meta-analyses of previous studies, provide the assessments of heterogeneity across these studies | NA | Not given by the studies from which summary data was acquired |
|  | d) | For two-sample MR:  i.  Provide justification of the similarity of the genetic variant-exposure associations between the exposure and outcome samples  ii.  Provide information on the number of individuals who overlap between the exposure and outcome studies | 21 |  |
| 11 | **Main results** |  |  |  |
|  | a) | Report the associations between genetic variant and exposure, and between genetic variant and outcome, preferably on an interpretable scale | NA |  |
|  | b) | Report MR estimates of the relationship between exposure and outcome, and the measures of uncertainty from the MR analysis, on an interpretable scale, such as odds ratio or relative risk per SD difference | Figure 4 and 5 and Extended data 4,10,11 |  |
|  | c) | If relevant, consider translating estimates of relative risk into absolute risk for a meaningful time period | NA |  |
|  | d) | Consider plots to visualize results (e.g. forest plot, scatterplot of associations between genetic variants and outcome versus between genetic variants and exposure) | Figure 4, 5 and Extended Data 11 |  |
| 12 | **Assessment of assumptions** |  |  |  |
|  | a) | Report the assessment of the validity of the assumptions | 8, 13, 14, Extended Data 4 |  |
|  | b) | Report any additional statistics (e.g., assessments of heterogeneity across genetic variants, such as *I^2^*, Q statistic or E-value) |  |  |
| 13 | **Sensitivity analyses and additional analyses** |  |  |  |
|  | a) | Report any sensitivity analyses to assess the robustness of the main results to violations of the assumptions | Extended Data 4, GitHub |  |
|  | b) | Report results from other sensitivity analyses or additional analyses | Extended Data 4, GitHub |  |
|  | c) | Report any assessment of direction of causal relationship (e.g., bidirectional MR) | Extended Data 4, GitHub |  |
|  | d) | When relevant, report and compare with estimates from non-MR analyses | NA |  |
|  | e) | Consider additional plots to visualize results (e.g., leave-one-out analyses) | GitHub |  |
|  | **DISCUSSION** |  |  |  |
| 14 | **Key results** | Summarize key results with reference to study objectives | 19 |  |
| 15 | **Limitations** | Discuss limitations of the study, taking into account the validity of the IV assumptions, other sources of potential bias, and imprecision. Discuss both direction and magnitude of any potential bias and any efforts to address them | 20,21 |  |
| 16 | **Interpretation** |  |  |  |
|  | a) | Meaning: Give a cautious overall interpretation of results in the context of their limitations and in comparison with other studies | 21 |  |
|  | b) | Mechanism: Discuss underlying biological mechanisms that could drive a potential causal relationship between the investigated exposure and the outcome, and whether the gene-environment equivalence assumption is reasonable. Use causal language carefully, clarifying that IV estimates may provide causal effects only under certain assumptions | 19,20 |  |
|  | c) | Clinical relevance: Discuss whether the results have clinical or public policy relevance, and to what extent they inform effect sizes of possible interventions | NA |  |
| 17 | **Generalizability** | Discuss the generalizability of the study results (a) to other populations, (b) across other exposure periods/timings, and (c) across other levels of exposure | NA |  |
|  | **OTHER INFORMATION** |  |  |  |
| 18 | **Funding** | Describe sources of funding and the role of funders in the present study and, if applicable, sources of funding for the databases and original study or studies on which the present study is based | 26 |  |
| 19 | **Data and data sharing** | Provide the data used to perform all analyses or report where and how the data can be accessed, and reference these sources in the article. Provide the statistical code needed to reproduce the results in the article, or report whether the code is publicly accessible and if so, where | 22 |  |
| 20 | **Conflicts of Interest** | All authors should declare all potential conflicts of interest | 26 |  |

This checklist is copyrighted by the Equator Network under the Creative Commons Attribution 3.0 Unported (CC BY 3.0) license.

1.	Skrivankova VW, Richmond RC, Woolf BAR, Yarmolinsky J, Davies NM, Swanson SA, et al. Strengthening the Reporting of Observational Studies in Epidemiology using Mendelian Randomization (STROBE-MR) Statement. JAMA. 2021;under review.

2.	Skrivankova VW, Richmond RC, Woolf BAR, Davies NM, Swanson SA, VanderWeele TJ, et al. Strengthening the Reporting of Observational Studies in Epidemiology using Mendelian Randomisation (STROBE-MR): Explanation and Elaboration. BMJ. 2021;375:n2233.
